# Female contraceptive cover duration for miltefosine-containing regimens for the treatment of women with leishmaniasis

**DOI:** 10.64898/2026.08.31.26360682

**Authors:** Wan-Yu Chu, Fabiana Alves, Thomas P.C. Dorlo

## Abstract

**Introduction:** Miltefosine is the only approved oral antileishmanial agent, but its use in women of childbearing potential (WOCBP) is restricted due to preclinical teratogenicity. Current labeling recommends contraception during treatment and for at least five months thereafter, based solely on its long terminal elimination half-life. This study re-evaluated the required contraceptive duration using an exposure margin-based approach.

**Methods:** Virtual populations were generated from anthropometric data of 382 Indian, 4,462 Eastern African, and 4,019 Brazilian WOCBP with leishmaniasis. Published population pharmacokinetic models were used to simulate miltefosine exposure following 14-42 days regimens for visceral leishmaniasis (VL), post-kala-azar dermal leishmaniasis (PKDL), and cutaneous leishmaniasis (CL). A developmental safety exposure threshold was derived from the rat no-observed-adverse-effect level (0.6 mg/kg/day for 10 days) and adjusted using a 10-fold safety margin. Contraceptive durations resulting in median residual post-contraception exposure (AUC_EOC-∞_) below this threshold were considered supportive of contraceptive discontinuation.

**Results:** The developmental safety exposure threshold was estimated at 2.5 mg·day/L. Despite pharmacokinetic differences across geographical regions and disease manifestations, required minimum contraceptive durations were consistent: three months from treatment initiation for the 14-day regimen, four months for 21- and 28-day regimens, and five months for the 42-day regimen. For the 14-day regimen, a single dose of a long-acting injectable contraceptive administered at treatment initiation would provide sufficient coverage.

**Conclusion:** An exposure margin-based approach supports shorter contraceptive durations than current recommendations. For the 14-day VL regimen, three months of contraception may provide a practical alternative to current labeling.

## 1. Introduction

Miltefosine is the first and only effective oral drug approved for the treatment of the neglected tropical parasitic infection leishmaniasis. Since its first registration in 2002 in India, miltefosine has been widely used for the treatment of visceral leishmaniasis (VL) in South Asia (as first-line therapy until 2014 and subsequently as part of combination regimens used as second-line therapy), for patients with VL and HIV co-infection in South Asia and Eastern Africa, for post-kala-azar dermal leishmaniasis (PKDL) in South Asia, and for cutaneous leishmaniasis (CL) in Latin America.^1,2^ Latest evidence on shorter miltefosine-containing combination regimens has supported updated treatment recommendations for patients with VL in Eastern Africa, and PKDL in both Eastern Africa and South Asia.^3^ Its oral administration, prolonged tissue residence time, and broad antiparasitic activity against various Leishmania species have made miltefosine a key component of global leishmaniasis control programmes.

Despite these advantages, the broader use of miltefosine, particularly among women of childbearing potential (WOCBP), has been limited by concerns about embryo-fetal toxicity. Preclinical studies demonstrated embryo- and fetotoxic effects in both rats and rabbits. Teratogenicity was observed only in rats, with drug-related malformations identified at the lowest observed adverse effect level (LOAEL) of 1.2 mg/kg/day following administration over a 10-day period during gestation.^4^ Consequently, miltefosine is contraindicated during pregnancy, and current product labelling recommends verifying pregnancy status prior to initiating miltefosine treatment (negative pregnancy test is required) and the use of effective contraception during treatment and for at least five months after the end of therapy.^5^

This recommendation is primarily based on miltefosine’s long terminal plasma elimination half-life (approximately 30 days), following the conventional assumption that near-complete drug clearance occurs after five elimination half-lives.^5^ While pragmatic, this half-life-based approach is largely theoretical and does not directly reflect the relationship between systemic exposure and developmental toxicity risk.^6^ Moreover, the terminal half-life is sensitive to sampling design and may not accurately represent the clinically relevant decline in exposure.^7^ Therefore, extrapolation based solely on half-life may be overly conservative, particularly for compounds with prolonged terminal phases such as miltefosine.^8^ Importantly, the prolonged contraception requirement has been shown to deter WOCBP from initiating miltefosine treatment or participating in clinical trials, thereby limiting access to therapy for leishmaniasis.^9^

Evolving regulatory guidance has introduced a more mechanistically relevant framework for developmental toxicity risk assessment. The ICH S5(R3) guideline emphasizes comparison of systemic exposure at the no observed adverse effect level (NOAEL) with clinical exposure, with increased concern when exposure margins are <10-fold and reduced or minimal concern at higher margins.^10^ Retrospective analyses of known human teratogens support this paradigm, showing that developmental toxicity typically occurs at exposures close to, or only a few-fold above, clinical levels, whereas larger margins are associated with low risk.^6^ Consistent with this, the European Federation of Pharmaceutical Industries and Association/PreClinical Development Expert Group (EFPIA-PDEG) consensus advocates integrating exposure margins into contraception guidance, while recognizing that the conventional five half-life rule is a pragmatic approximation rather than a biologically grounded threshold.^11^ Together, these frameworks support an exposure margin-based approach as a more quantitative and clinically relevant basis for assessing developmental toxicity risk and informing contraception recommendations.

In light of the recently released updates to WHO treatment guidelines for VL and PKDL,^3^ including shorter miltefosine-containing regimens (e.g., 14 days for VL combination therapy and 21 or 42 days for PKDL depending on region), it is timely to re-evaluate the current contraception recommendations using an exposure margin-based approach. Given the well-characterized population pharmacokinetics of miltefosine across diverse patient populations, this study aimed to re-assess the minimally required duration of contraceptive cover following miltefosine treatment by integrating preclinical developmental toxicity data with human population pharmacokinetic simulations informed by anthropometric data from historical VL cohorts in India and Eastern Africa, as well as CL cohorts from Brazil. By accounting for known determinants of exposure, including body size, disease manifestations, and geographic region, the objective was to derive estimates of contraceptive duration for different treatment regimens and populations that ensure developmental safety while minimizing unnecessary barriers to treatment access for WOCBP.

## 2. Methods

### 2.1. Demographic data and virtual populations

Demographic data for VL patients were compiled from multiple sources: datasets from Médecins Sans Frontières (MSF) collected in Lankien, South Sudan between 1998 and 2019; pharmacovigilance data from Drugs for Neglected Diseases initiative (DNDi) trials conducted in Ethiopia, Kenya, Sudan, and Uganda;^12^ and datasets from the MSF Operational Centre Barcelona-Athens collected between 2007 and 2009 at Hajipur SADR Hospital in Vaishali District, Bihar State, India. For CL patients, demographic data were obtained from DATASUS/SINAN, the Brazilian Ministry of Health’s national disease surveillance database.

Women of childbearing age (15-49 years) with available body weight (WT), height, and age data were extracted from the database. Body mass index (BMI) and fat-free mass (FFM) were calculated, with FFM derived using a sex-specific, age-including equation for females (Equation 1):^13^

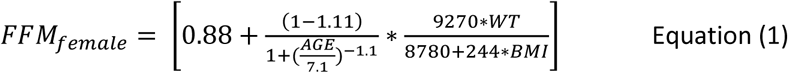

For the Brazilian database, height data were unavailable, precluding calculation of BMI and use of Equation 1. Therefore, FFM was estimated as WT × (1 − body fat fraction), with body fat fraction sampled from a normal distribution (mean=0.359, SD=0.094) based on published data from Brazilian young adults.^14^

Using these data, virtual populations of 10,000 Eastern African, 10,000 Indian and 10,000 Brazilian people with leishmaniasis were simulated using a conditional distribution modeling approach, in which correlated covariates were generated sequentially conditional on previously simulated variables.^15^ Conditional relationships were estimated from the original data, preserving empirical correlations between covariates while allowing extrapolation beyond the observed population for clinical trial simulations. Age was used as the seeding variable for the imputation process and was sampled from the original dataset.

### 2.2. Population pharmacokinetics model

Individual plasma concentration-time profiles of miltefosine were simulated using previously developed population pharmacokinetic models, with minor adjustments (*Supplementary Material S1 and S2*). Simulations for VL and PKDL were based on models developed using data from people with VL and PKDL in Eastern African,^16^ and simulations for CL were based on a model developed using data from people with CL from Colombia,^17^ The final model parameters applied in the simulations are summarized in *Supplementary Table S1 and S2*. All simulations were conducted using the “mrgsolve” package in R.^18^

### 2.3. Simulated miltefosine regimens

Simulations were based on miltefosine regimens previously evaluated in clinical studies conducted in Eastern Africa, South Asia, and Latin America. Treatment durations of 14 and 28 days were simulated for VL,^19^ 21, 28, and 42 days for PKDL,^20,21^ and 21 and 28 days for CL.^22^ For patients weighting < 30 kg, miltefosine was dosed according to the weight-band-based regimen recommended in the updated WHO guidelines (*Supplementary Table S3*).^3,23^ For patients weighing ≥30 kg, dosing followed the conventional regimen, with patients weighing 30-44 kg receiving 100 mg/day and those weighing ≥45 kg receiving 150 mg/day.

### 2.4. Definition of developmental safety exposure threshold in human

In accordance with the ICH S5 (R3) guideline on the detection of reproductive and developmental toxicity for human pharmaceuticals,^10^ the developmental risk assessment was based on comparison of systemic drug exposure at the NOAEL in the most sensitive animal species with human exposure, using the most relevant exposure metric.

In embryo-fetal development toxicity studies, oral administration of miltefosine at doses of ≥1.2 mg/kg/day for ten consecutive days during gestation in female rats resulted in teratogenic effects. The NOAEL for developmental toxicity was therefore established at 0.6 mg/kg/day.^4^

For miltefosine, the area under the plasma concentration-time curve (AUC) was selected as the primary exposure metric, as it reflects cumulative systemic exposure during prolonged oral treatment and is appropriate given the drug’s long elimination half-lives. Systemic exposure at the NOAEL was defined as the area under the concentration-time curve from time zero to infinity (AUC_0-∞_), calculated as:

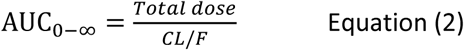

The PK characteristics of miltefosine in rats have been described in the literature. Available PK parameter estimates indicate that exposure increases approximately dose-proportionally over the 1-10 mg/kg range, with median apparent clearance (CL/F) in male rats reported to range between 0.2 and 0.5 L/day/kg.^4,24^ In the absence of PK data at the NOAEL, extrapolation of AUC from doses within a 3-fold range of the NOAEL is considered toxicologically acceptable;^6^ therefore, CL/F values reported in the literature were used to derive AUC at the NOAEL. Although sex-specific clearance estimates in female rats are not directly available, repeated-dose studies suggest that AUC in female rats were approximately 1.03- to 1.10-fold higher than in males.^4^

The female rat AUC_0-∞_ at the NOAEL was taken as the basis for defining the developmental safety exposure threshold. In line with ICH S5 (R3) guidance, which indicates increased concern when the NOAEL occurs at exposures less than 10-fold higher than human exposure, this value was conservatively divided by a factor of ten to define the exposure limit used for developmental risk classification.^10^ Human exposures exceeding this adjusted threshold were considered indicative of a potential developmental toxicity risk.

### 2.5. Assessment of residual exposure after contraceptive coverage

The unprotected residual exposure to miltefosine after the end of the contraceptive cover period until infinity (AUC_EOC-∞_; schematically illustrated in **Figure 1**) was quantified from individual simulated pharmacokinetic profiles in virtual Eastern African, Indian, and Brazilian WOCBP with VL, PKDL, and CL using Equation 3:

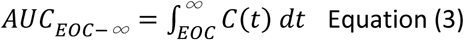

**Figure 1.**
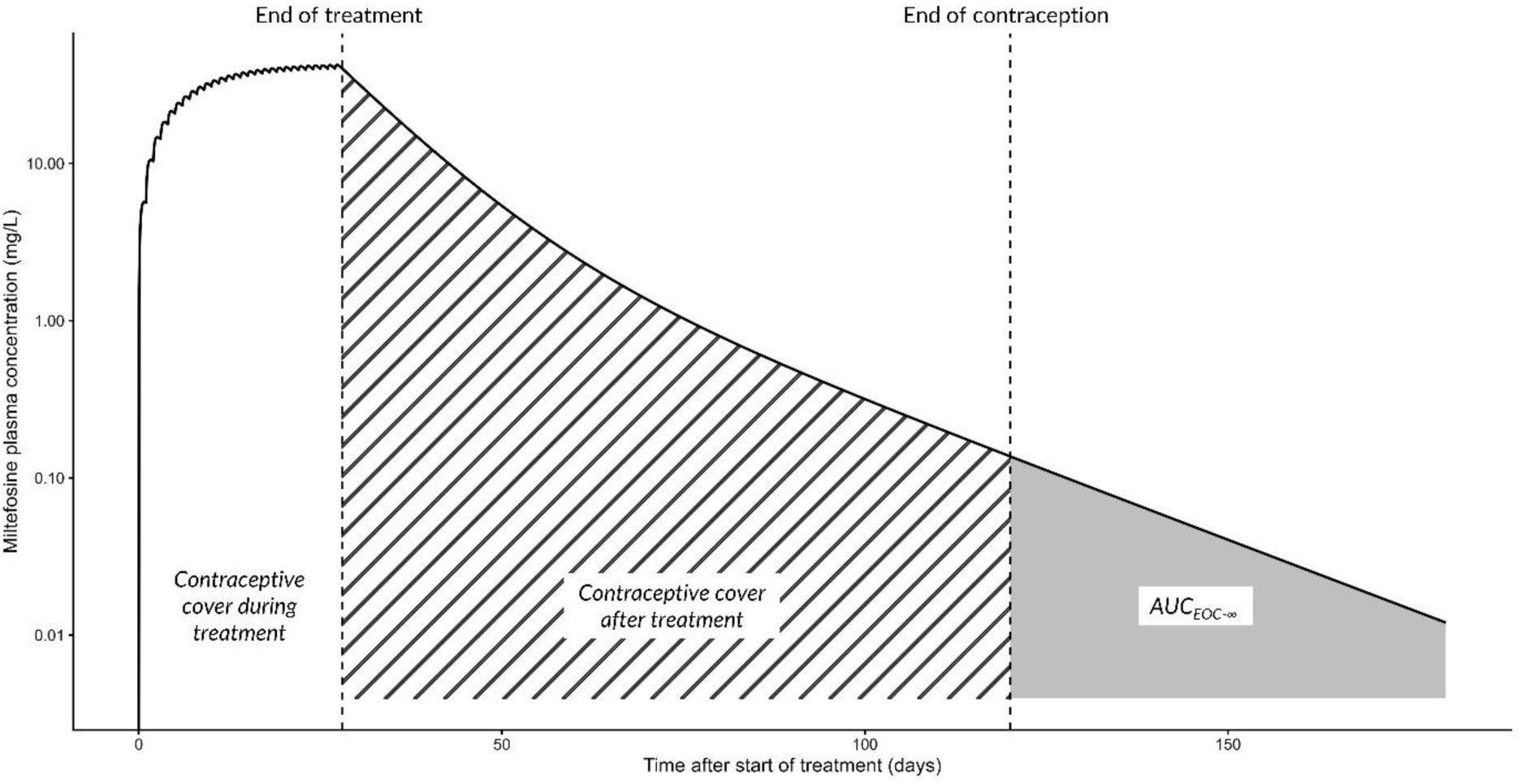
Schematic depiction of contraceptive cover in relation to miltefosine exposure. *AUC_EOC-∞_ denotes unprotected residual exposure to miltefosine after the end of the contraceptive cover period until infinity*.

Analyses were performed across the evaluated miltefosine treatment durations: 14 and 28 days for VL, 21, 28, and 42 days for PKDL, and 21 and 28 days for CL.

Consistent with ICH M3 (R2) guideline (general toxicity studies), safety margins are typically evaluated using central exposure metrics (e.g., group mean or median AUC) rather than individual extremes.^25^ Furthermore, in accordance with ICH S5 (R3) guideline, developmental safety margins of ≥ 10-fold are generally considered to represent a reduced clinical concern and are an acceptable threshold for risk assessment.^10^ Applying these principles to the population simulations, scenarios in which the median exposure of the simulated patient populations remained below the predefined developmental safety exposure threshold (*Section 2.4*) were considered scientifically supportive of discontinuing contraception.

Contraceptive coverage periods of one to six months from the start of treatment were evaluated, corresponding to one or two doses of depot medroxyprogesterone acetate (DMPA), which provides approximately three months of contraceptive protection per dose. This reflects typical trial practice, in which participants receive one dose prior to treatment initiation and a second dose at the 3- month follow-up visit.

## 3. Results

### 3.1 Demographic data and virtual populations

The demographic datasets comprised 382 Indian and 4,462 Eastern African WOCBP with VL or PKDL, and 4,019 Brazilian WOCBP with CL. Their demographic characteristics are summarized in **Table 1**. The Eastern African demographics were predominantly derived from VL cohorts in South Sudan (>95%). Populations from South Sudan are largely of Nilotic ancestry and, on average, exhibit greater height and a leaner body habitus compared with neighboring populations in Sudan, Ethiopia, Kenya, and Uganda, which are genetically heterogeneous.

**Table 1.** Demographic characteristics of women of childbearing potential.

|  | Indian (VL) | Eastern Africa,<br>excl. South<br>Sudan (VL) | South Sudan (VL) | Brazil (CL) |
| --- | --- | --- | --- | --- |
|  | (n=382) | (n = 201) | (n=4261) | (n=4019) |
| **Age (years)** | 26 (20, 35) | 21 (17, 28) | 25 (18, 31) | 34 (24, 41) |
| **Weight (kg)** | 39 (35, 44) | 44 (40, 49) | 47 (42, 52) | 67 (58, 78) |
| **Height (cm)** | 149 (145, 152) | 161 (157, 165) | 173 (167, 179) | - |
| **BMI (kg/m<sup>2</sup>)** | 17.6 (16.2, 19.0) | 16.7 (15.5, 18.5) | 15.7 (14.4, 17.0) | - |
| **Fat-free mass (kg)** | 29 (26, 31) | 32 (30, 35) | 35 (32, 38) | 42 (36, 51) <sup>a</sup> |
| **Country composition, n (%)** | - | Ethiopia 26 (13%)<br>Kenya 63 (31%)<br>Sudan 99 (49%)<br>Uganda 13 (7%) | - | - |

Data are presented as median (interquartile range [IQR]) unless otherwise indicated.
VL, visceral leishmaniasis; CL, cutaneous leishmaniasis.
<sup>a</sup> Fat-free mass was calculated as body weight × (1 – body fat fraction), with the body fat fraction sampled from a normal distribution (mean=0.359, SD=0.094) derived from published data in Brazilian young adults.<sup>14</sup>

To mitigate potential bias resulting from the dominance of South Sudanese individuals, a simulation-based resampling approach was applied. Specifically, a virtual population of 6,000 females representing Eastern African populations excluding South Sudan was generated using conditional anthropometric distributions derived from the available data (n=201). This virtual population was combined with the South Sudan dataset (n = 4,261) to construct a resampled Eastern African population of 10,000 individuals. The Indian and Brazilian populations were independently generated using an analogous simulation-based expansion approach, yielding 10,000 virtual individuals per population. Demographic characteristics of the virtual populations are summarized in *Supplementary Table S4*, which closely resemble those of the original patient populations.

### 3.2. Population pharmacokinetic simulation

In the combined virtual population of 30,000 individuals, only 27 individuals (<0.1%, 24 from India and 4 from Eastern Africa) had a body weight between 25 and 30 kg and therefore received the 80 mg/day miltefosine dose. In the Indian VL/PKDL virtual population, 86% of individuals received 100 mg/day and 14% received 150 mg/day, whereas in the Eastern African VL/PKDL virtual population, 47% and 53% received 100 mg/day and 150 mg/day, respectively, reflecting the smaller body size of the South Asian population. In the Brazilian CL virtual population, 98% of individuals received the 150 mg/day dose and the remaining 2% received 100 mg/day. This distribution reflects the generally better nutritional status of CL patients compared with VL/PKDL patients. Furthermore, the CL population had an older age distribution than the VL/PKDL populations, which likely contributed to the higher body weights and corresponding dose allocation.

For each individual, miltefosine concentration-time profiles were simulated based on disease-specific pharmacokinetic characteristics for VL, PKDL and CL, with known geographical differences incorporated. Treatment durations of 14 and 28 days were simulated for VL, 21, 28, and 42 days for PKDL, and 21 and 28 days for CL. **Figure 2** presents the median concentrations during and after treatment together with the 95% prediction intervals for the respective dose regimens. **Table 2** summarizes the simulated AUC_0-∞_ values by treatment regimen, disease manifestation, and geographical region.

**Figure 2.**
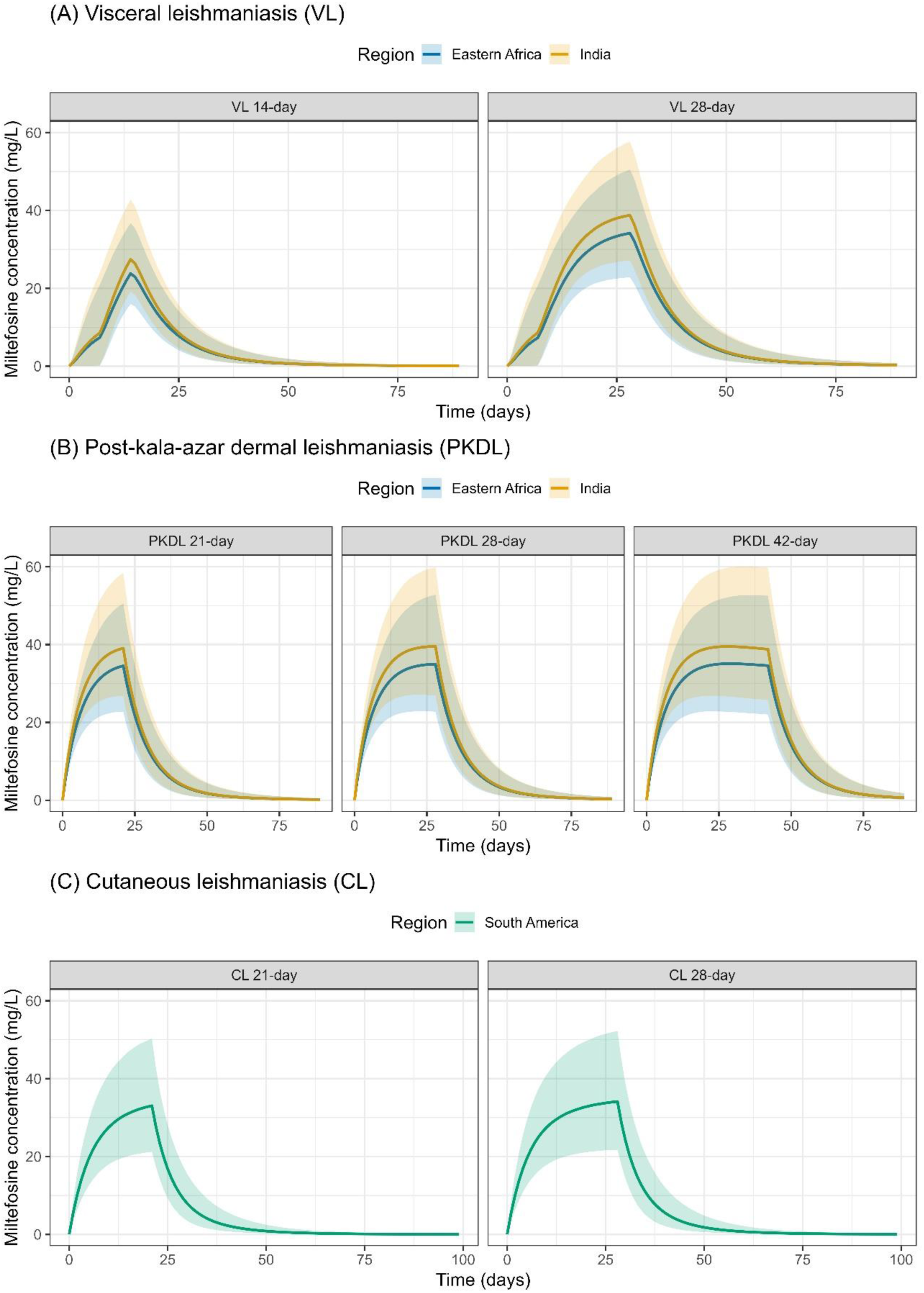
Simulated miltefosine plasma concentration-time profiles in A) patients with visceral leishmaniasis (VL) receiving 14-day and 28-day regimens, B) patients with post-kala-azar leishmaniasis (PKDL) receiving 21-day, 28-day, and 42-day regimens and C) patients with cutaneous leishmaniasis (CL) receiving 21-day and 28-day regimens. *The solid line represents the population median, and the shaded area represents the 95% prediction interval*.

**Table 2.** Simulated miltefosine AUCs (mg/L·day) for various treatment durations stratified by population and regimen.

| Population/<br>regimen | Simulated miltefosine AUCs (mg/L·day) |  |  |  |
| --- | --- | --- | --- | --- |
|  | 14-day | 21-day | 28-day | 42-day |
| **VL Eastern Africa** | 388<br>(306, 490) | - | 929<br>(789, 1102) | - |
| **VL India** | 438<br>(346, 553) | - | 1055<br>(900, 1236) | - |
| **PKDL Eastern Africa** | - | 894<br>(762, 1044) | 1156<br>(989, 1352) | 1671<br>(1426, 1947) |
| **PKDL India** | - | 1006<br>(875, 1157) | 1305<br>(1134, 1511) | 1875<br>(1626, 2168) |
| **CL Latin America** | - | 800<br>(678, 936) | 1064<br>(911, 1243) | - |

Data are presented as median (interquartile range [IQR]) unless otherwise indicated.
VL, visceral leishmaniasis; CL, cutaneous leishmaniasis; PKDL, post-kala-azar dermal leishmaniasis.

### 3.3. Developmental safety exposure threshold

Based on literature-reported pharmacokinetic parameters, systemic exposure at the NOAEL (0.6 mg/kg/day administered for 10 days, cumulative dose of 6.0 mg/kg) was derived assuming dose-proportional pharmacokinetics.^4^ A median CL/F of 0.26 L/day/kg in male rats was used, while CL/F in female rats was assumed to be 0.24 L/day/kg, reflecting a 1.10-fold higher systemic exposure.^4^ Using the female clearance estimate, the AUC_0-∞_ at the NOAEL was calculated to be 25 mg·day/L. Applying a default 10-fold safety margin, a conservative developmental safety exposure threshold of 2.5 mg·day/L was defined.

### 3.4. Assessment of residual exposure after contraceptive coverage and minimum required contraceptive durations

Simulated individual concentration-time profiles were generated for each miltefosine dosing regimen in VL and PKDL populations from India and Eastern Africa, and in a CL population from Brazil, from which individual AUC_EOC-∞_ values were derived. Contraceptive duration requirements did not differ between Eastern African and Indian patients, nor between VL and PKDL patients receiving the same treatment duration (*Supplementary Material S5*). **Table 3** summarizes the median AUC_EOC-∞_ values for contraception durations of one to six months after treatment initiation stratified by disease and treatment durations. Scenarios in which the median AUC_EOC-∞_ fell below the developmental safety exposure of 2.5 mg·day/L were considered supportive of discontinuing contraception.

**Table 3.**
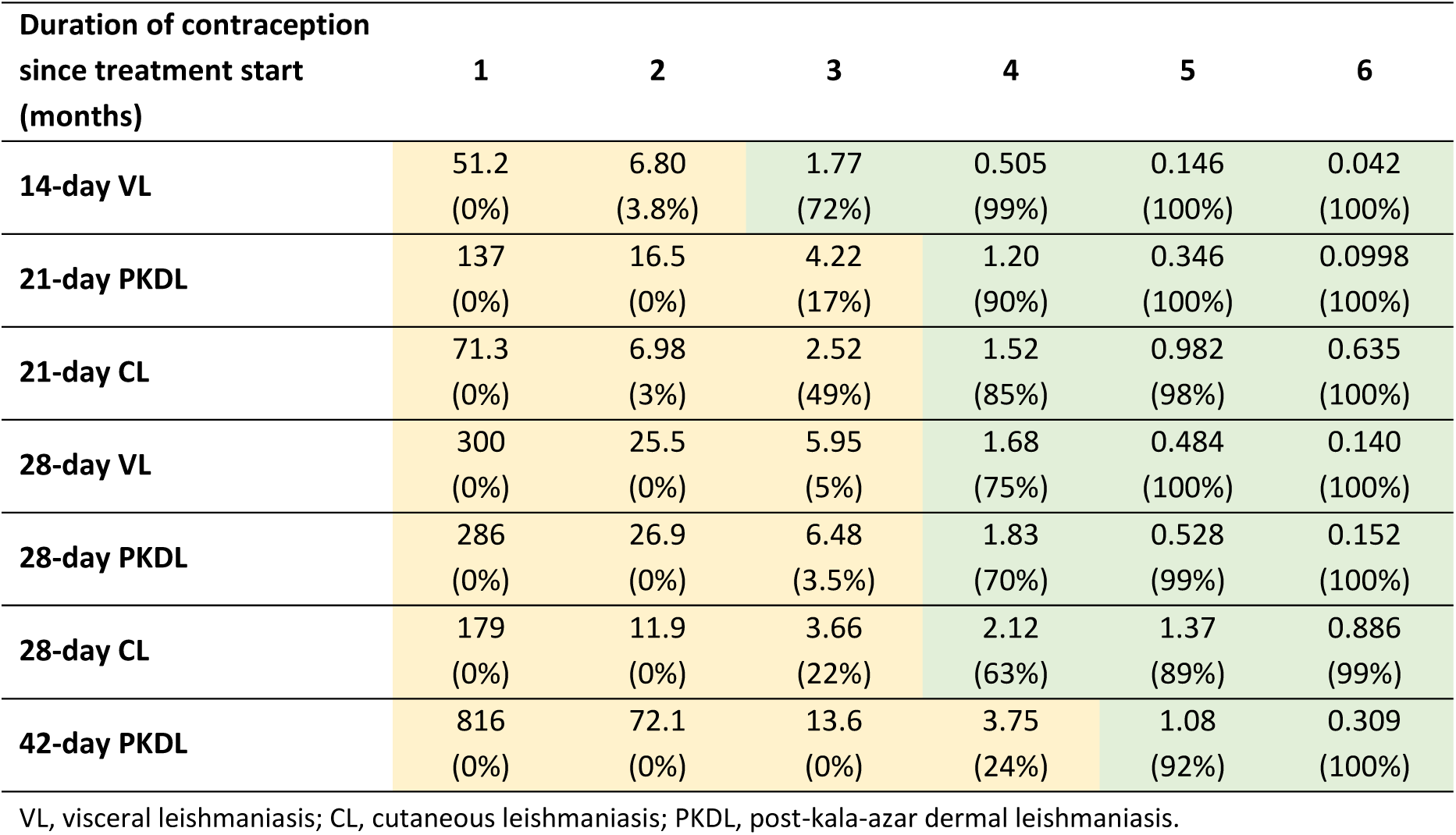
Median AUC_EOC-∞_ (mg/L·day) below the developmental safety exposure threshold. *AUC_EOC-∞_ denotes unprotected residual exposure to miltefosine after the end of the contraceptive cover period until infinity. The developmental safety threshold was defined as 2.5 mg·day/L. Medians above or equal the developmental safety threshold are shaded yellow. Medians below the developmental safety threshold are shaded green. Values in parentheses indicate the percentage of individuals with* AUC_EOC-∞_ *below the developmental safety exposure threshold*.

| Duration of contraception since treatment start (months) | 1 | 2 | 3 | 4 | 5 | 6 |
| --- | --- | --- | --- | --- | --- | --- |
| 14-day VL | 51.2<br>(0%) | 6.80<br>(3.8%) | 1.77<br>(72%) | 0.505<br>(99%) | 0.146<br>(100%) | 0.042<br>(100%) |
| 21-day PKDL | 137<br>(0%) | 16.5<br>(0%) | 4.22<br>(17%) | 1.20<br>(90%) | 0.346<br>(100%) | 0.0998<br>(100%) |
| 21-day CL | 71.3<br>(0%) | 6.98<br>(3%) | 2.52<br>(49%) | 1.52<br>(85%) | 0.982<br>(98%) | 0.635<br>(100%) |
| 28-day VL | 300<br>(0%) | 25.5<br>(0%) | 5.95<br>(5%) | 1.68<br>(75%) | 0.484<br>(100%) | 0.140<br>(100%) |
| 28-day PKDL | 286<br>(0%) | 26.9<br>(0%) | 6.48<br>(3.5%) | 1.83<br>(70%) | 0.528<br>(99%) | 0.152<br>(100%) |
| 28-day CL | 179<br>(0%) | 11.9<br>(0%) | 3.66<br>(22%) | 2.12<br>(63%) | 1.37<br>(89%) | 0.886<br>(99%) |
| 42-day PKDL | 816<br>(0%) | 72.1<br>(0%) | 13.6<br>(0%) | 3.75<br>(24%) | 1.08<br>(92%) | 0.309<br>(100%) |

VL, visceral leishmaniasis; CL, cutaneous leishmaniasis; PKDL, post-kala-azar dermal leishmaniasis.

A comparison of the minimally required total contraceptive durations derived using the half-life-based and exposure margin-based approach is presented in **Table 4**. The table also outlines potential contraceptive strategies for meeting the exposure margin-based requirements, including DMPA administered alone or in combination with an additional oral contraceptive method (e.g., a one-month course corresponding to a standard pill cycle, assuming correct daily use).

**Table 4.** Minimally required total contraceptive durations and potential contraceptive strategies for various durations of miltefosine therapy.

| Approach | Recommended minimum total contraceptive duration derived using different approaches <sup>a</sup> |  |  |  |
| --- | --- | --- | --- | --- |
|  | 14-day | 21-day | 28-day | 42-day |
| Half-life based <sup>5</sup> | 5.5 months | 6 months | 6 months | 6.5 months |
| Dose-extrapolation <sup>8</sup> | 2.5 months <sup>b</sup> | - | 5 months | - |
| Exposure margin-based | 3 months | 4 months | 4 months | 5 months |
| **Contraceptive strategies to meet the minimum duration for <u>exposure margin-based results</u>** |  |  |  |  |
| Long-acting injectable DMPA | 1 dose | 2 doses | 2 doses | 2 doses |
| Long-acting injectable DMPA + oral contraception | 1 dose DMPA | 1 dose DMPA + 1 month oral | 1 dose DMPA + 1 month oral | 1 dose DMPA + 2 month oral |

DMPA, depot medroxyprogesterone acetate, providing contraceptive protection for 3 months.
<sup>a</sup> Duration of contraception since treatment start.
<sup>b</sup> Recommendations for a 14-day miltefosine regimen were not provided; values refer to simulations of a 10-day treatment regimen.<sup>8</sup>

The shortest contraceptive duration requirement was observed for the 14-day miltefosine-containing VL regimen. Three months of contraception from treatment initiation was sufficient for the median AUC_EOC-∞_ to fall below the developmental safety exposure threshold in both Eastern African and Indian populations (*Supplementary Figure S1*), meeting the guideline-defined safety criterion. This requirement could be readily covered by a single dose of injectable DMPA administered at treatment initiation.

For 21- and 28-day miltefosine-containing regimens, a minimum total contraceptive duration of four months from treatment initiation was required for the median AUC_EOC-∞_ to fall below the developmental safety exposure threshold. This recommendation was consistent across VL, PKDL and CL populations and geographical regions.

For the 42-day regimen used in PKDL, a total contraceptive duration of five months from treatment initiation was required, with approximately 90% of simulated individuals predicted to have AUC_EOC-∞_ values below the predefined developmental safety threshold.

## 4. Discussion

Recently released WHO treatment guidelines for VL and PKDL incorporate evidence from recent clinical trials supporting shorter treatment regimens,^3^ including a 14-day miltefosine-paromomycin combination as an effective and more patient-friendly treatment option for VL.^19^ However specific contraceptive guidance for a 14-day regimen is lacking. Current WHO recommendations advise effective contraception during treatment and for 2 months after treatment for shorter miltefosine regimens (≤10 days), or for 5 months after treatment for regimens of ≥28 days.^3^ The current product label recommends contraception during treatment and for 5 months after the last dose regardless of treatment duration, resulting in approximately 5.5 months of contraceptive coverage when applied to a 14-day regimen.^5^ This lack of specific guidance creates uncertainty regarding the appropriate contraceptive duration for a 14-day regimen, while applying the prolonged product-label recommendation may pose a major barrier to its broader implementation in women. The present study provides a refined estimate, suggesting that 3 months of contraceptive coverage from treatment initiation is sufficient for a 14-day miltefosine-containing regimen. This period can be readily covered by a single dose of a long-acting injectable contraceptive administered at treatment start, potentially removing an important operational barrier to implementing the miltefosine-paromomycin regimen in routine VL care.

The current miltefosine product-label recommendation follows the traditional half-life-based approach, with the post-treatment contraceptive period derived from its long terminal elimination half-life.^5^ A subsequent dose-extrapolation approach sought to refine this recommendation by accounting for differences in dose and treatment duration.^8^ However, this approach relied on converting the animal NOAEL to a human equivalent dose and indirectly estimating the corresponding human exposure, introducing assumptions in the translation from dose to systemic exposure. In the present study, we further advanced this assessment using an exposure margin-based framework that directly compares predicted human systemic exposure with exposure at the NOAEL in preclinical species. This approach is consistent with ICH-aligned toxicity risk assessment principles, which emphasize systemic exposure for evaluating human safety margins.^10,11^ Direct comparison of systemic exposures reduces reliance on dose-based extrapolation and better accounts for differences in miltefosine exposure across treatment regimens and populations, providing a more pharmacologically informed and clinically relevant basis for determining the required contraceptive duration.

In this study, pharmacokinetic differences related to body size, disease manifestation (VL, PKDL, and CL), and geographical region (Eastern Africa, South Asia, Latin America) were explicitly accounted for. Despite these differences, the required contraception duration depended mainly on regimen length and was consistent across populations. From the start of miltefosine treatment, a total contraception duration of three months was sufficient for 14-day regimens, four months was sufficient for 21- and 28-day regimens, and five months was sufficient for the 42-day regimen, to ensure post-treatment exposure remains below the developmental safety threshold. This corresponds to an approximate reduction of two months compared with current label recommendations based on the half-life approach, and a reduction of one month for the 28-day treatment regimens compared with previous estimates derived using the dose-extrapolation approach (**Table 4**).

In routine leishmaniasis case management, contraception options are discussed with the patient to ensure selection of the most appropriate method, given their current and past use and their preferences. Due to the potential gastro-intestinal side effects of miltefosine (nausea and vomiting are common in the first week(s) of therapy), an injectable long-acting contraceptive (e.g., DMPA) providing reliable coverage for three months would be a preferable option to oral contraceptives. When the contraception recommendation is longer than three months, the patient may opt for a second long-lasting injection, requiring a visit to the clinic at three months after treatment onset, or opt for an extension of the contraceptive therapy with other methods, depending on their preference (e.g., oral contraceptives). In practice, DMPA is a highly effective injectable contraceptive with good individual-level acceptability in South Asia, Eastern Africa, and Latin America, particularly when supported by appropriate counseling.^27–29^ This pragmatic approach minimizes reliance on prolonged adherence while remaining feasible in routine clinical settings, although effective counselling (to the women and extended to their partners) remains important to ensure reliable contraceptive coverage during the recommended period.

This study demonstrates that developmental safety following miltefosine treatment can be ensured with substantially shorter and more pragmatic contraceptive protection than the currently recommended period, particularly for the 14-day miltefosine-paromomycin regimen. An exposure margin-based, model-informed framework provides a robust basis for refining these recommendations. Importantly, simplifying and shortening contraception requirements addresses a key operational access barrier for miltefosine in WOCBP and supports improved acceptability, equity, and feasibility of miltefosine-containing regimens in routine care and public health programmes.

## Data Availability

All data produced in the present study are available upon reasonable request to the authors.

## Acknowledgement

We gratefully acknowledge Médecins Sans Frontières (MSF), the Drugs for Neglected Diseases initiative (DNDi), and DATASUS, Sistema de Informação de Agravos de Notificação (SINAN), Secretaria de Vigilância em Saúde e Ambiente (SVSA) do Ministério da Saúde, Brasil, for providing access to the demographic datasets used in this study.

## Transparency declarations

None to declare.

## References

1. Pareyn M, Alves F, Burza S, et al. Leishmaniasis. Nat Rev Dis Primer 2025; 11: 81.

2. World Health Organization. WHO guideline for the treatment of visceral leishmaniasis in HIV co-infected patients in East Africa and South-East Asia. 2022. Available at: https://www.who.int/publications/i/item/9789240048294. Accessed May 29, 2026.

3. World Health Organization. WHO guidelines on leishmaniases: treatment of visceral leishmaniasis and post-kala-azar dermal leishmaniasis in eastern Africa and South-East Asia. 2026. Available at: https://www.who.int/publications/i/item/9789240123298. Accessed August 16, 2026.

4. Wild JS. Pharmacology/toxicology NDA/BLA review and evaluation: miltefosine (Impavido), NDA 204684. U.S. Food and Drug Administration, Center for Drug Evaluation and Research; 2013. Available at: https://www.accessdata.fda.gov/drugsatfda_docs/nda/2014/204684Orig1s000PharmR.pdf. Accessed April 7, 2026.

5. Paladin Therapeutics Inc. IMPAVIDO (miltefosine) capsules, for oral use: Prescribing Information. U.S. Food and Drug Administration; 2014. Available at: https://www.accessdata.fda.gov/drugsatfda_docs/label/2014/204684s000lbl.pdf. Accessed January 15, 2026.

6. Andrews PA, Blanset D, Costa PL, et al. Analysis of exposure margins in developmental toxicity studies for detection of human teratogens. Regul Toxicol Pharmacol RTP 2019; 105: 62–8.

7. Toutain PL, Bousquet-Mélou A. Plasma terminal half-life. J Vet Pharmacol Ther 2004; 27: 427–39.

8. Dorlo TPC, Balasegaram M, Lima MA, de Vries PJ, Beijnen JH, Huitema ADR. Translational pharmacokinetic modelling and simulation for the assessment of duration of contraceptive use after treatment with miltefosine. J Antimicrob Chemother 2012; 67: 1996–2004.

9. Sunyoto T, Potet J, Boelaert M. Why miltefosine-a life-saving drug for leishmaniasis-is unavailable to people who need it the most. BMJ Glob Health 2018; 3: e000709.

10. International Council for Harmonisation of Technical Requirements for Pharmaceuticals for Human Use. ICH S5(R3) guideline on detection of reproductive and developmental toxicity for human pharmaceuticals. 2020. Available at: https://www.ema.europa.eu/en/ich-s5-r3-guideline-detection-reproductive-developmental-toxicity-human-pharmaceuticals-scientific-guideline. Accessed January 15, 2026.

11. Bowman CJ, Becourt-Lhote N, Boulifard V, et al. Science-Based Approach to Harmonize Contraception Recommendations in Clinical Trials and Pharmaceutical Labels. Clin Pharmacol Ther 2023; 113: 226–45.

12. Kimutai R, Musa AM, Njoroge S, et al. Safety and Effectiveness of Sodium Stibogluconate and Paromomycin Combination for the Treatment of Visceral Leishmaniasis in Eastern Africa: Results from a Pharmacovigilance Programme. Clin Drug Investig 2017; 37: 259–72.

13. Al-Sallami HS, Goulding A, Grant A, Taylor R, Holford N, Duffull SB. Prediction of Fat-Free Mass in Children. Clin Pharmacokinet 2015; 54: 1169–78.

14. Branco BHM, Bernuci MP, Marques DC, et al. Proposal of a normative table for body fat percentages of Brazilian young adults through bioimpedanciometry. J Exerc Rehabil 2018; 14: 974–9.

15. Smania G, Jonsson EN. Conditional distribution modeling as an alternative method for covariates simulation: Comparison with joint multivariate normal and bootstrap techniques. CPT Pharmacomet Syst Pharmacol 2021; 10: 330–9.

16. Chu W-Y, Verrest L, Younis BM, et al. Disease-Specific Differences in Pharmacokinetics of Paromomycin and Miltefosine Between Post-Kala-Azar Dermal Leishmaniasis and Visceral Leishmaniasis Patients in Eastern Africa. J Infect Dis 2024; 230: e1375–84.

17. Kip AE, Castro M del M, Gomez MA, et al. Simultaneous population pharmacokinetic modelling of plasma and intracellular PBMC miltefosine concentrations in New World cutaneous leishmaniasis and exploration of exposure–response relationships. J Antimicrob Chemother 2018; 73: 2104–11.

18. Baron KT. mrgsolve: Simulate from ODE-Based Models. 2026. Available at: https://CRAN.R-project.org/package=mrgsolve. Accessed January 15, 2026.

19. Musa AM, Mbui J, Mohammed R, et al. Paromomycin and Miltefosine Combination as an Alternative to Treat Patients With Visceral Leishmaniasis in Eastern Africa: A Randomized, Controlled, Multicountry Trial. Clin Infect Dis Off Publ Infect Dis Soc Am 2023; 76: e1177–85.

20. Sundar S, Pandey K, Mondal D, et al. A phase II, non-comparative randomised trial of two treatments involving liposomal amphotericin B and miltefosine for post-kala-azar dermal leishmaniasis in India and Bangladesh. PLoS Negl Trop Dis 2024; 18: e0012242.

21. Younis BM, Mudawi Musa A, Monnerat S, et al. Safety and efficacy of paromomycin/miltefosine/liposomal amphotericin B combinations for the treatment of post-kala-azar dermal leishmaniasis in Sudan: A phase II, open label, randomized, parallel arm study. PLoS Negl Trop Dis 2023; 17: e0011780.

22. López L, Valencia B, Alvarez F, et al. A phase II multicenter randomized study to evaluate the safety and efficacy of combining thermotherapy and a short course of miltefosine for the treatment of uncomplicated cutaneous leishmaniasis in the New World. PLoS Negl Trop Dis 2022; 16: e0010238.

23. Mazariegos Herrera A, Karlsson MO, Svensson EM, Dorlo TPC. Weight-band-based simplification of oral allometric miltefosine dosing in paediatric patients with visceral leishmaniasis. J Antimicrob Chemother 2026; 81: dkag014.

24. Valicherla GR, Tripathi P, Singh SK, et al. Pharmacokinetics and bioavailability assessment of Miltefosine in rats using high performance liquid chromatography tandem mass spectrometry. J Chromatogr B Analyt Technol Biomed Life Sci 2016; 1031: 123–30.

25. International Council for Harmonisation of Technical Requirements for Pharmaceuticals fo. ICH M3(R2) guideline on non-clinical safety studies for the conduct of human clinical trials and marketing authorisation for pharmaceuticals – Step 5. 2013. Available at: https://www.ema.europa.eu/en/ich-m3-r2-non-clinical-safety-studies-conduct-human-clinical-trials-pharmaceuticals-scientific-guideline. Accessed January 15, 2026.

26. Dorlo TPC, Balasegaram M, Beijnen JH, de Vries PJ. Miltefosine: a review of its pharmacology and therapeutic efficacy in the treatment of leishmaniasis. J Antimicrob Chemother 2012; 67: 2576–97.

27. Ayuk BE, Yankam BM, Saah FI, Bain LE. Provision of injectable contraceptives by community health workers in sub-Saharan Africa: a systematic review of safety, acceptability and effectiveness. Hum Resour Health 2022; 20: 66.

28. Srivastava Garg S, Bhateja B, Grover S, Tapasvi I. Injection Depot Medroxyprogesterone Acetate as an Early Postpartum Contraceptive Measure: Evaluation of Its Acceptability, Efficacy, and Impact on Lactation. Cureus 16: e66454.

29. Léo Persoz et al. Exploring gender-inclusive approaches to improve Chagas disease & leishmaniasis treatment access for women (ASTMH 2025 Annual Meeting; Abstract #7336). Available at: https://d1lx8xdon3af7k.cloudfront.net/ASTMH_2025_Annual_Meeting_Abstract_Book_56833b3855.pdf.

