## Supplementary Material for "Female contraceptive cover duration for miltefosine-containing regimens for the treatment of women with leishmaniasis"

#### S1. Miltefosine population pharmacokinetic model in VL and PKDL

The miltefosine pharmacokinetic models used for simulations in patients with visceral Leishmaniasis (VL) and post-kala-azar dermal leishmaniasis (PKDL) were based on published population pharmacokinetic models developed using data from Eastern African populations.[1] The structural model consisted of a two-compartment disposition model with first-order absorption and first-order elimination from the central compartment.

To account for variability in body size, fat-free mass (FFM) was used as the body size descriptor. In the original models, FFM was incorporated through allometric scaling of apparent clearance (CL/F) and the central volume of distribution (V2/F), with fixed exponents of 0.75 and 1, respectively. In the present study, allometric scaling was extended to all clearance parameters, including intercompartmental clearance (Q), and all volume parameters, including both central and peripheral volumes. The models were subsequently re-estimated, with all clearance and volume parameters normalized to an FFM of 53 kg. Final parameter estimates are provided in **Tables S1**.

In addition to body size effects, disease-related and regional differences in miltefosine pharmacokinetics identified in previous studies were incorporated into the simulations. In VL patients, a reduction in relative oral bioavailability (F) during the first week of treatment was included to account for disease-related malabsorption. Furthermore, a cumulative miltefosine dose effect on F was incorporated in both VL and PKDL patients.[1] Previous studies have also reported geographical differences in miltefosine pharmacokinetics, with higher overall drug exposure observed in South Asian patients than in Eastern African patients with VL and PKDL.[2] In PKDL, this difference was quantified as an approximately 18% higher relative bioavailability in South Asian patients and was incorporated into the simulations accordingly.[3]

**Table S1**. Final miltefosine pharmacokinetic model parameters in VL and PKDL.

| Parameter | Unit | Estimate |
|---|---|---|
| **Absorption rate in PKDL ($k_a$ PKDL)** | 1/day | 5.15 |
| **Absorption rate in VL ($k_a$ VL)** | 1/day | 0.85 |
| **Clearance [a] (CL/F)** | L/day | 2.9 |
| **Volume of central compartment [a] (V2/F)** | L | 29 |
| **Inter-compartmental clearance [a] (Q/F)** | L/h | 0.163 |
| **Volume of peripheral compartment [a] (V3/F)** | L | 4.2 |
| **Relative change in F during the first week of treatment in VL patients ($COV_{F,VL}$)** | - | 0.67 |
| **Exponent of power relationship between cumulative MF dose and F ($COV_{F,CD}$)** | - | -0.11 |
| **Percentage increase in F in Indian patients compared with Eastern African patients ($COV_{F,GEO}$)** | - | 18 |
| **Between subject variability in CL/F** | CV% | 19 |
| **Between subject variability in V2/F** | CV% | 11[b] |
| **Between subject variability in $COV_{F,VL}$** | CV% | 70 |

*VL, visceral Leishmaniasis; PKDL, post-kala-azar dermal leishmaniasis.*

[a] Allometric scaled based on fat-free mass with power exponent of 0.75 for clearance and 1 for volume of distribution. Estimate is given for a standardized fat free mass of 53 kg.

[b] Between subject variability in V2/F was not unidentifiable, but high correlation between CL/F and V2/F was suggested by SIR result. Therefore, the same η distribution was assumed for CL/F and V2/F, and a scaling factor of 0.37 was estimated.

### S2. Miltefosine population pharmacokinetic model in CL.

Miltefosine pharmacokinetic model used for simulations in patients with cutaneous leishmaniasis (CL) was based on previously published population pharmacokinetic models developed in Colombian population without adjustment.[4] The structural model consisted of a three-compartment disposition model with first-order absorption and first-order elimination from the central compartment. Final parameter estimates are provided in **Tables S2**.

**Table S2**. Final miltefosine pharmacokinetic model parameters in CL.

| Parameter | Unit | Estimate |
|---|---|---|
| **Absorption rate in CL ($k_a$ CL)** | 1/day | 9.6 |
| **Clearance [a] (CL/F)** | L/day | 4.62 |
| **Volume of central compartment [a] (V2/F)** | L | 28.5 |
| **Inter-compartmental clearance (Q3/F)** | L/day | 0.42 |
| **Volume of peripheral compartment (V3/F)** | L | 3.85 |
| **Inter-compartmental clearance (Q4/F)** | L/day | 0.0274 |
| **Volume of peripheral compartment (V4/F)** | L | 2.02 |
| **Between subject variability in CL/F** | CV% | 15 |
| **Between subject variability in V2/F** | CV% | 11 |

[a] Allometric scaled based on fat-free mass with power exponent of 0.75 for clearance and 1 for volume of distribution. Estimate is given for a standardized fat free mass of 53 kg.

### S3. Miltefosine dosing regimen

**Table S3**. WHO-recommended weight-band regimen for patients weighing <30 kg.[5]

| Weight band range (kg) | Daily dose (mg) |
|---|---|
| **<6** | 20 |
| **6.00-9.99** | 30 |
| **10.00-14.99** | 50 |
| **15.00-19.99** | 60 |
| **20.00-24.99** | 70 |
| **25.00-29.99** | 80 |

### S4. Simulated virtual populations

**Table S4**. Demographic characteristics of virtual populations

| | Indian VL/PKDL (n=10,000) | Eastern African VL/PKDL (n=10,000) | Brazilian CL (n=10,000) |
|---|---|---|---|
| **Age (years)** | 25<br>[20, 35] | 23<br>[17, 30] | 33<br>[24, 41] |
| **Weight (kg)** | 39<br>[36, 42] | 45<br>[40, 49] | 67<br>[58, 77] |
| **Height (cm)** | 150<br>[142, 156] | 166<br>[157, 175] | - |
| **BMI (kg/m²)** | 17.4<br>[16.2, 19.0] | 16.0<br>[14.8, 17.5] | - |
| **Fat-free mass (kg)** | 28<br>[26, 30] | 33<br>[30, 37] | 42<br>[36, 50] |
| **Country (%)** | - | Ethiopia 7.5%<br>Kenya 19%<br>Sudan 28%<br>Uganda 3.5%<br>South Sudan 42% | - |

Data are presented as median (interquartile range [IQR]) unless otherwise indicated.
VL, visceral Leishmaniasis; PKDL, post-kala-azar dermal leishmaniasis; CL, cutaneous leishmaniasis.

### S5. 95% prediction intervals of $AUC_{EOC-\infty}$ and the proportion of patients with $AUC_{EOC-\infty}$ values below the developmental safety threshold.

**Figure S1.** Post-contraceptive miltefosine exposure ($AUC_{EOC-\infty}$) compared with the developmental safety exposure threshold (median and 95% prediction interval). *The dashed line denotes exposure at the NOAEL (25 mg·day/L), and the solid line represents the developmental safety threshold (2.5 mg·day/L), derived using a default 10-fold safety margin. Red percentages indicate the proportion of patients below the developmental safety threshold.*

*(A)* Visceral Leishmaniasis (VL)

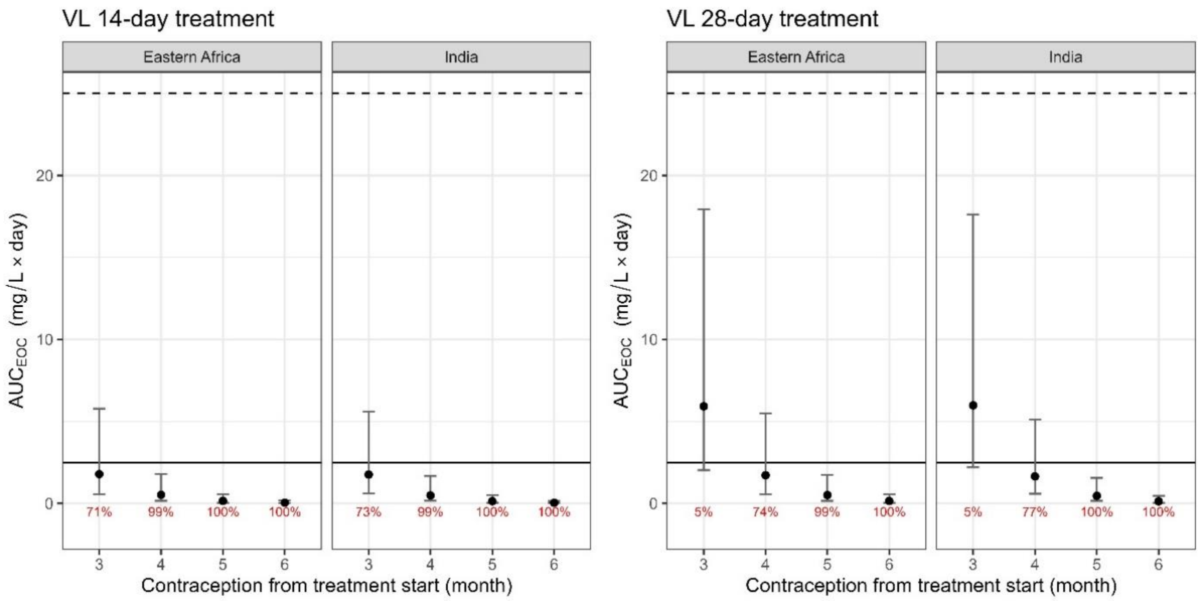

(B) Post-kala-azar dermal leishmaniasis (PKDL)

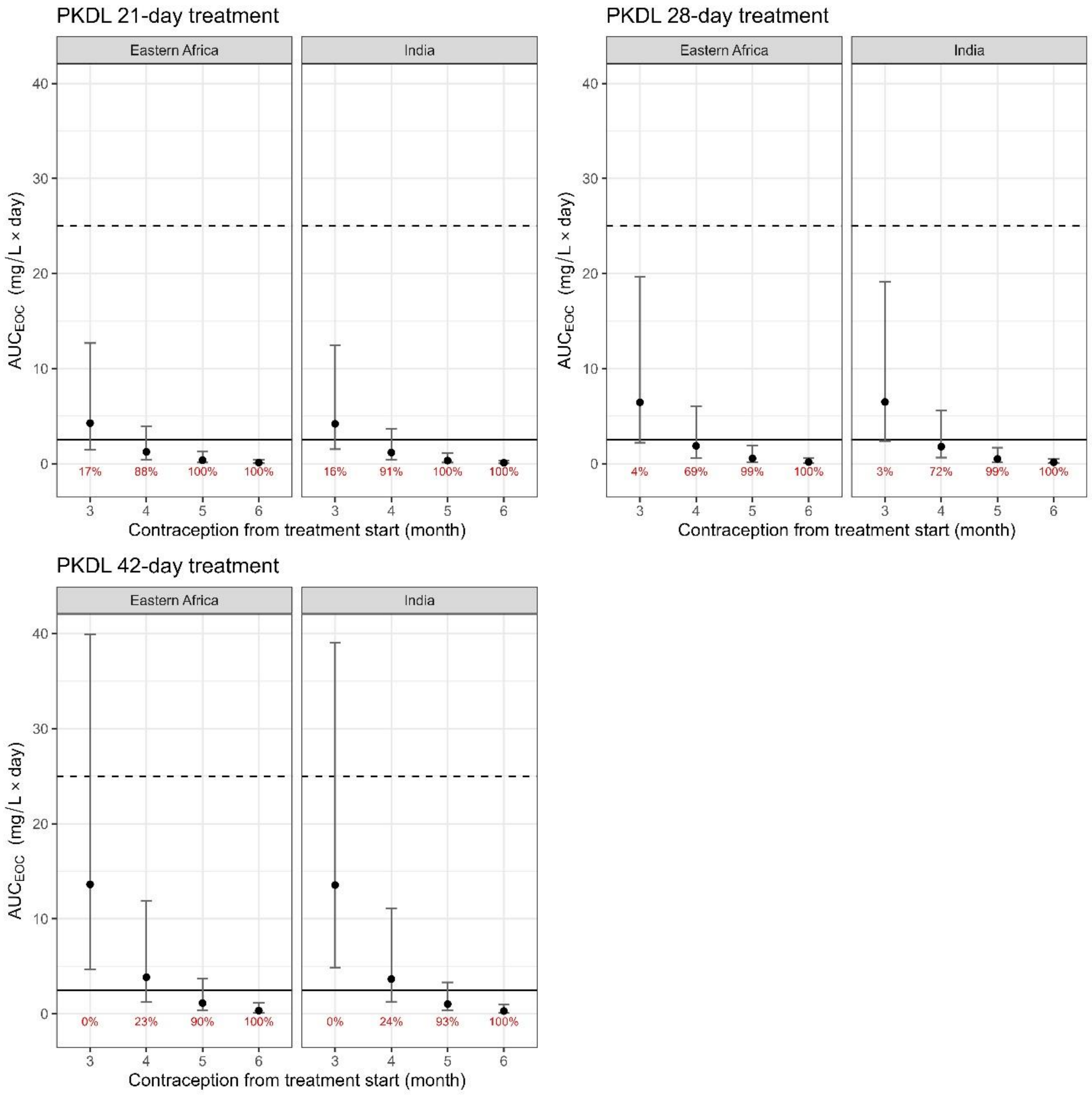


(C) Cutaneous leishmaniasis (CL)

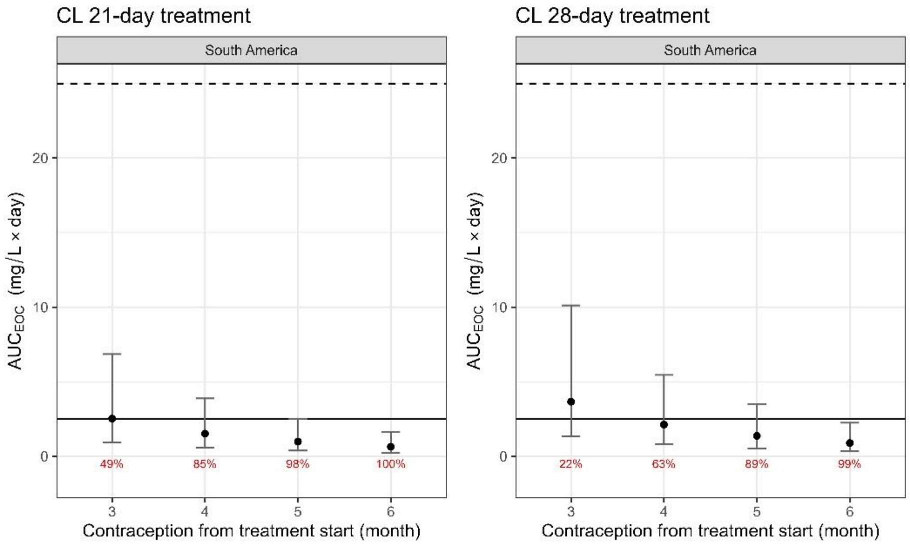

### References


1. Chu W-Y, Verrest L, Younis BM, *et al.* Disease-Specific Differences in Pharmacokinetics of Paromomycin and Miltefosine Between Post-Kala-Azar Dermal Leishmaniasis and Visceral Leishmaniasis Patients in Eastern Africa. *J Infect Dis* 2024; **230**: e1375–84.

2. Dorlo TPC, Balasegaram M, Beijnen JH, de Vries PJ. Miltefosine: a review of its pharmacology and therapeutic efficacy in the treatment of leishmaniasis. *J Antimicrob Chemother* 2012; **67**: 2576–97.

3. Chu W-Y. Advancing Drug Therapy in Poverty-Related Diseases. 2025. Available at: https://dspace.library.uu.nl/items/4a84297e-b2a1-45c5-910b-1c1afb670006. Accessed June 23, 2026.

4. Kip AE, Castro M del M, Gomez MA, *et al.* Simultaneous population pharmacokinetic modelling of plasma and intracellular PBMC miltefosine concentrations in New World cutaneous leishmaniasis and exploration of exposure–response relationships. *J Antimicrob Chemother* 2018; **73**: 2104–11.

5. Mazariegos Herrera A, Karlsson MO, Svensson EM, Dorlo TPC. Weight-band-based simplification of oral allometric miltefosine dosing in paediatric patients with visceral leishmaniasis. *J Antimicrob Chemother* 2026; **81**: dkag014.
